# Evoked response signatures explain deep brain stimulation outcomes

**DOI:** 10.1101/2024.10.04.24314308

**Authors:** Bahne H. Bahners, Lukas L. Goede, Garance M. Meyer, Luise Poser, Lauren A. Hart, Julianna Pijar, Nanditha Rajamani, Barbara Hollunder, Savir Madan, Simón Oxenford, Gunnar Waterstraat, Gabriel Curio, Alfons Schnitzler, Esther Florin, Andrea A. Kühn, Michael D. Fox, Andreas Horn

## Abstract

Deep brain stimulation (DBS) is an established treatment for Parkinson’s disease. Still, DBS parameter programming currently follows a tedious trial-and-error process. DBS-evoked cortical potentials (EP) might guide parameter selection but this concept has not yet been tested. Further, mounting *wet* EEG systems is too time-consuming to scale in outpatient clinic settings. Here, we test the utility of a novel method that leverages the *spatial pattern* of EP using a *dry* EEG setup. We acquired EP in 58 hemispheres in patients with Parkinson’s disease and compute a model which represents the optimal EP response pattern associated with maximal clinical improvements. Once defined, we use this pattern to estimate stimulation outcomes in unseen patients. Finally, we utilize it to identify optimal stimulation contacts in five unseen hemispheres where it selected the correct contact in all cases. The simple setup makes this novel method an attractive option to guide DBS programming in clinical practice.

## Introduction

Deep brain stimulation (DBS) of the subthalamic nucleus (STN) is an effective therapy for patients with Parkinson’s disease^1^. DBS devices allow distributing electric currents in 1% steps across multiple DBS contacts, leading to a large number of ∼10^10^ options for cathodal current distribution on an eight-contact directional DBS electrode ^2^. Even when following heuristic strategies that sample the parameter space in a smart fashion, the current process of selecting optimal stimulation contacts requires multiple permutations through different stimulation parameters along with clinical testing ^3,4^. This process is time-consuming and inconvenient for both patients and healthcare providers. Hence, novel strategies to guide DBS programming are needed ^5,6^. One promising strategy, termed image-guided programming, is to localize DBS electrodes and define optimal stimulation targets ^7^. Based on such electrode reconstructions, an algorithm may identify the parameter settings that maximize stimulating the identified ‘sweet-spot’ ^8^ or optimal circuit ^9^. This approach is promising and has been successful in a first prospective clinical trial^10^. However, one potential shortcoming is that the approach does not measure or capture target engagement. To address this gap, *Boutet et al.* proposed to measure optimal DBS response profiles using fMRI^5^. Here, individual patients were scanned under multiple stimulation settings to determine an optimal response profile that could be used to optimize their DBS parameters. While promising and scientifically interesting, the approach requires MRI scanning time, which involves high cost and the need for data processing expertise which may preclude this approach from becoming practical in a global clinical setting.

Cortical potentials evoked by STN-DBS (EP) have been investigated in electroencephalography (EEG) recordings as early as the first clinical DBS trials ^11–14^. The concept here is to measure the cortical response of individual DBS pulses, i.e., cortical responses are averaged time-locked to independent pulses delivered at the electrode level. Recent studies using magnetoencephalography (MEG) and EEG, observed that the amplitudes of DBS-EP over the motor cortex and the supplementary motor area scale with motor performance and the therapeutic window ^6,15,16^. With this understanding in mind, EP, and especially their amplitudes over the motor cortex, have been proposed as a tool to guide DBS programming ^15–19^. Still, while the concept is a logical next step, none of these published studies have yet tested clinical applicability of such an EP-guided programming approach. Partly, this might result from the same issues as in the use of fMRI: scarce availability, technical difficulty, and high cost of fMRI equally apply to recording MEG during active DBS ^5,20,21^. Recording DBS-EP with conventional scalp EEG may instead be a more practical approach regarding cost and expertise. However, the time-consuming process of mounting gel electrodes, which often takes up to an hour for a skilled team and can be inconvenient for the patient ^22^, may still render this approach impractical for use at scale in typical clinical settings. In contrast, *dry* electrode EEG systems can be set up in minutes, without the need for highly skilled personnel. We explore this option in the present manuscript.

A second limitation of published EP-based frameworks is that these suggested to use EP *amplitudes of single channels* or brain regions to measure the optimality of DBS settings ^6,15,16^. For instance, the amplitude of the EP over the primary motor cortex would typically correlate with clinical improvements ^6,16^, making it a viable candidate for direct use in DBS programming. While promising, EP amplitudes and topographies may change considerably over time ^6^, are dependent on impedance of measurements and may hence vary from day to day and may differ across patients. Also, there is no agreement on which exact EP-latency or EEG channel should be used to extract EP amplitudes for DBS programming. Along the lines of DBS network mapping that used fMRI or diffusion MRI data ^23^, here, we propose to use the *global pattern of EP*, which aims at characterizing the spatial distribution – or network engaged by the stimulation across channels and EP time course. In this approach, amplitudes in neighboring channels (and amplitudes as distributed across the entire cortex) would normalize themselves, and it is more the pattern or distribution of EPs, rather than the single EP amplitude, that is used to inform optimality of DBS settings.

Here, we developed a strategy that leverages both the use of dry-EEG, which has the key strength in its ease-of-use, and the application of a global response pattern model, which may have the strength of robustness and transferability across patients. We call this approach **DBS**-**P**rogramming **U**sing **L**ightweight **S**etup **E**EG (*DBS-PULSE*). To test it, we recorded potentials evoked by STN stimulation in 58 hemispheres of patients with PD (discovery cohort) using a 32-channel dry-electrode setup. The approach correlated each element of the EP-map with clinical improvements across the patient cohort. A resulting correlation matrix then serves as a model of the optimal response and can be used to estimate motor improvements in unseen data points / novel patients. Indeed, following similar approaches we introduced in the field of connectomic DBS ^23^, this pattern aims at approximating the optimal EP distribution across *space and time*. We first subjected the pattern to cross-validation within the discovery cohort to test its potential generalizability and estimate degree of over-fitting to the training sample. Then, we used the pattern in an independent dataset. Namely, an independent prospective test cohort was acquired, recording dry-EEG EPs from a total of 20 DBS contacts in five additional hemispheres. In each patient, we applied the pattern to select the best contact that was predicted to yield best clinical improvements.

## Results

### Patient demographics and clinical results

We recruited a total of 32 patients (11 females; mean age: 60.4 ± 6.8 years) with Parkinson’s disease. All patients had received bilateral STN-DBS implantation (see Supplementary Table 1 for electrode models and stimulation settings) and there was a significant overall response to stimulation in the upper limb unified Parkinson’s disease rating scale part III items (UPDRS-III; items 3.3-3.6; 3.15-3.17; stimulation OFF: 18.9 ± 4.3 vs. stimulation ON: 10.2 ± 4.7; t (29) = 9.51, p = 0.001). We focused on the upper limb scores, given that in many centers upper limb symptoms are exactly what is being measured during monopolar review testing. During clinical testing and dry EEG recordings all patients were in their best medication on-state (mean levodopa equivalent daily dose, LEDD: 606 ± 276 mg). This was a purposeful and deliberate choice, since, if successful, the model would render more practical to implement in clinical routine. Indeed, patients typically visit an outpatient department in the medication on state, so we aimed at creating a system that would be suitable for this condition. The discovery cohort for primary analysis consisted of the first 29 patients (58 hemispheres) recruited for this study. Another three patients (data collected from five hemispheres, one hemisphere excluded due to fatigue, see Methods), that were not included in the discovery cohort, were recruited upon completing the primary analysis for prospective validation (test cohort) and underwent more extensive recordings (see below). A comprehensive summary of clinical characteristics, medication and demographics is provided in supplementary table 1. DBS electrode localization confirmed accurate placement of leads in the subthalamic nucleus for all patients included in the study (see supplementary figure S1). Figure 1 summarizes the methodological workflow.

**Figure 1.**
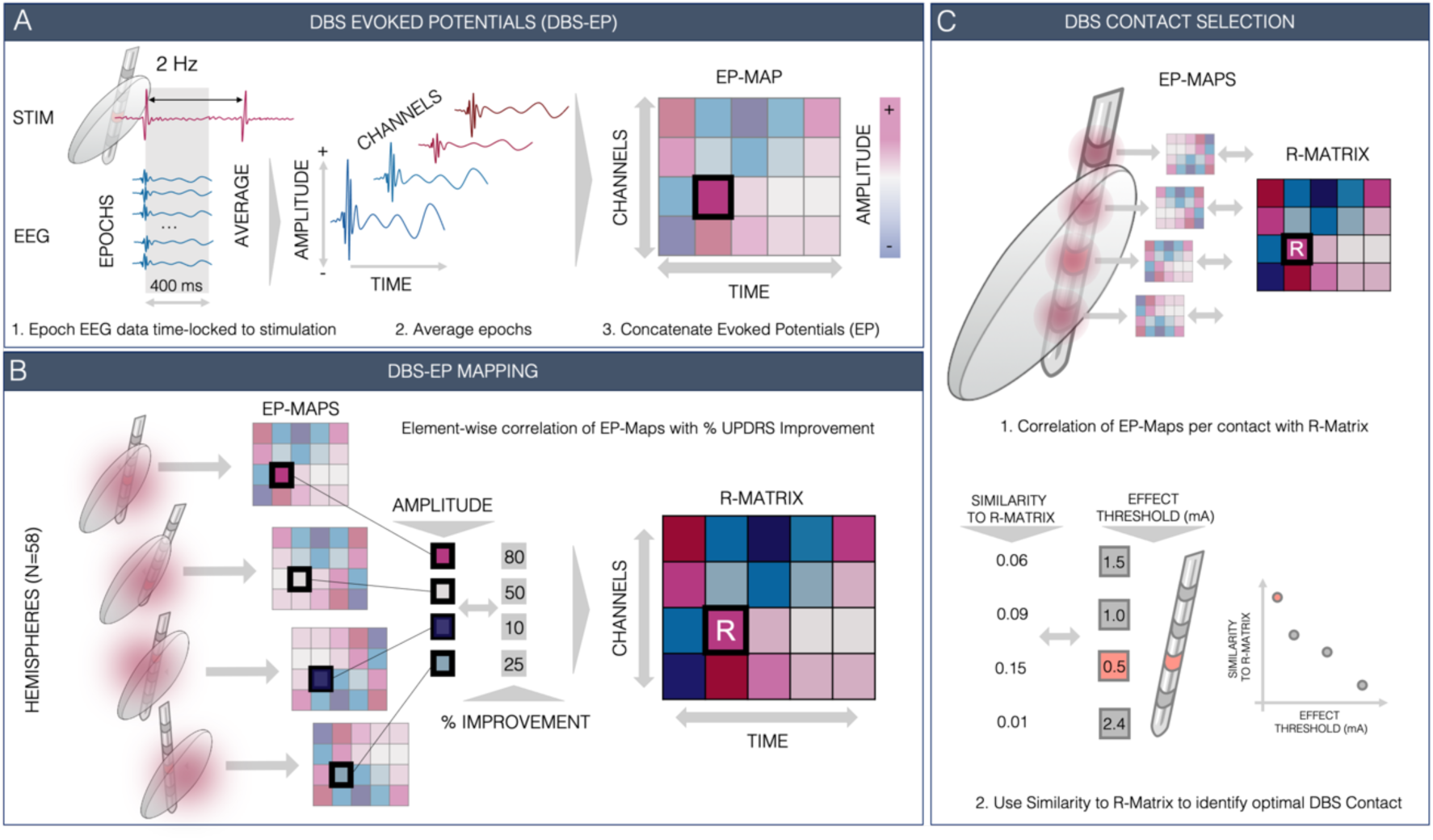
DBS-Evoked Potential Mapping. **A:** Stimulation pulses were applied with a frequency of 2 Hz. The data was divided into epochs (i.e. time segments) time-locked to the stimulation pulses. These epochs were averaged to obtain Evoked Potentials (EP) across channels. The EP amplitudes were then concatenated into a 32 channel × time matrix (EP-map). **B:** EP-maps resulting from stimulation in 58 hemispheres were used to apply an element-wise correlation between amplitude values of the EP-maps across hemispheres and contralateral Unified Parkinson’s disease Rating Scale part III (UPDRS-III) upper limb hemibody improvements. The resulting correlation matrix (R-matrix) may embody an optimal EP pattern. To test the validity of this pattern, it was used to estimate individual improvements in unseen patients (multiple cross-validation designs). To do so, (unseen) EP-maps were correlated (element-wise) with the R-Matrix, which led to a correlation coefficient that expresses agreement between the EP-map and the R-Matrix. This coefficient was correlated with clinical improvements. **C:** Application of the *DBS-PULSE* approach: In a prospective cohort, the same procedure was carried out for EP-maps after stimulating different contacts in the same patient. The resulting correlation coefficients (i.e. similarities to the R-matrix) were then used to identify the optimal deep brain stimulation (DBS) contact in these independent patients.

### DBS-EP reveal fronto-central topography

In the discovery cohort we applied a total of 57,503 unilateral subthalamic low-frequency (2 Hz) stimulation pulses (Fig. 2 C). The stimulation was applied at the chronic DBS contact using a monopolar stimulation montage in 58 hemispheres during dry EEG recordings (see supplementary table 1 for chronic stimulation settings). When using monopolar DBS during electrophysiological recordings, there is a characteristic decay artifact after the DBS pulse that may obscure short latency EP ^6,20^. To assess the amount of time after stimulation that was obscured by the DBS pulse artifact in our recordings, we developed a novel DBS-EEG phantom (supplementary figure S2). Based on the shape and length of the stimulation decay artifact in the phantom data, the earliest EP that could be distinguished from the artifact had a peak latency of around 20 ms (supplementary figure S2D). We therefore focused our analysis on long-latency EP (> 20 ms) that were unaffected by the stimulation artifact ^6,14^.

**Figure 2.**
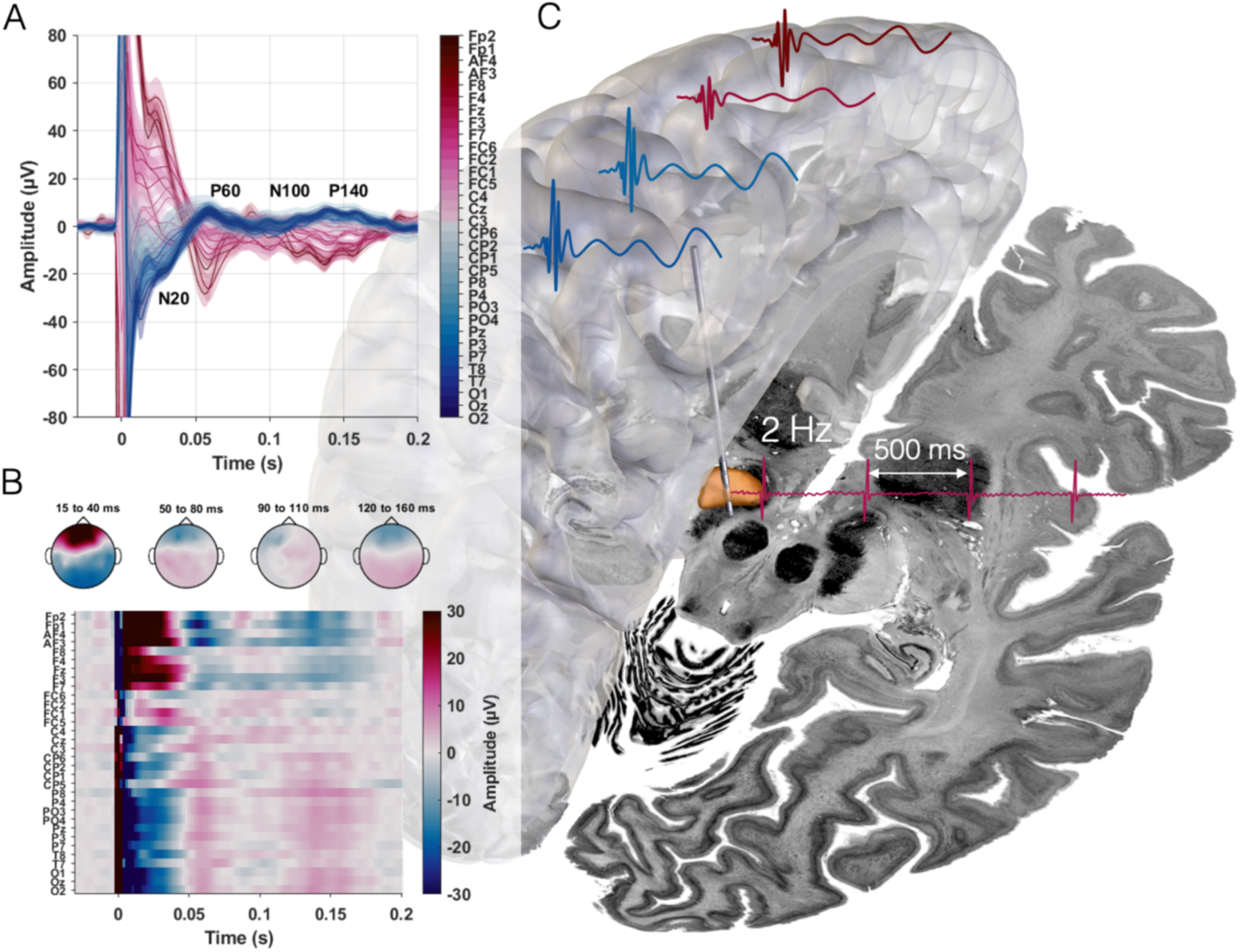
DBS-EP. **A:** Grand average deep brain stimulation-evoked potential (DBS-EP) time series of all 32 channels and standard error of the mean (shaded areas) across 58 hemispheres. In panels A and B channels are ordered along a fronto-occipital gradient. Potentials are labeled in reference to the central channels dominating polarity (N20 for the first negative peak in central channels at 20 ms, N100 for the negative peak in central channels at 100 ms, P60 and P140 for the positive peaks at 60 and 140 ms). B: Grand average EP map (channel x time) and topography plots averaged across the respective time windows of N20, P60, N100 and P140. C: Schematic visualization of DBS-EP generated with 2 Hz stimulation of the subthalamic nucleus (STN, shown in orange) across ipsilateral central and frontal cortical areas. The Big Brain template and DISTAL atlases were used as a backdrop image and for STN rendering respectively ^24,25^. The EP overlayed on top of a transparent volumetric rendering of a standardized cortical surface template in ICBM 2009b Nonlinear Asymmetric MNI space and the stimulation pulses next to the STN are schematic drawings.

The dry EEG data was epoched time-locked to the stimulation pulses, averaged across epochs and concatenated into EP-maps (Fig. 1A). Across hemispheres, stimulation evoked four distinct potentials at 20, 60, 100 and 140 ms after stimulation (referred to as N20, P60, N100 and P140 in the following, see Fig. 2 A). The N20 was characterized by a bipolar pattern with a frontal vs. parieto-central gradient and a left preponderance, while the bipolar parieto-frontal topographies of P60 and P140 were less lateralized. The N100 topography and amplitudes appeared least consistent across recordings (Fig. 2). Next, we aimed to compute a model that represents the optimal EP pattern an effective electrode should ideally elicit to induce maximal symptom relief. Hence, for each element of the EP-map, amplitudes were correlated with contralateral upper limb percentage UPDRS-III improvements obtained for the same contact under high-frequency DBS across 58 hemispheres, leading to a matrix of correlation coefficients (R-matrix) (Fig. 1 B). The topography of the resulting R-matrix showed three main clusters of clinically relevant responses (Fig. 3). First, the N20 pattern formed a positively correlated band across central EEG channels. Second, for P60 there was a bipolar pattern with parieto-occipital channel amplitudes negatively and frontal channels positively correlated with motor improvements. Third, the amplitude of parietal and central channels around 100 ms was positively correlated with clinical improvements (Fig. 3). Importantly, the 100 ms response only became prominent after correlation with clinical improvements and was less saliently observed in the grand average (Fig. 2B and 3A). Last, the amplitudes of median central and frontal channels around the P140 peak negatively correlated with improvements, while left-lateralized central channel amplitudes (FC5 and C3) showed a positive correlation.

**Figure 3.**
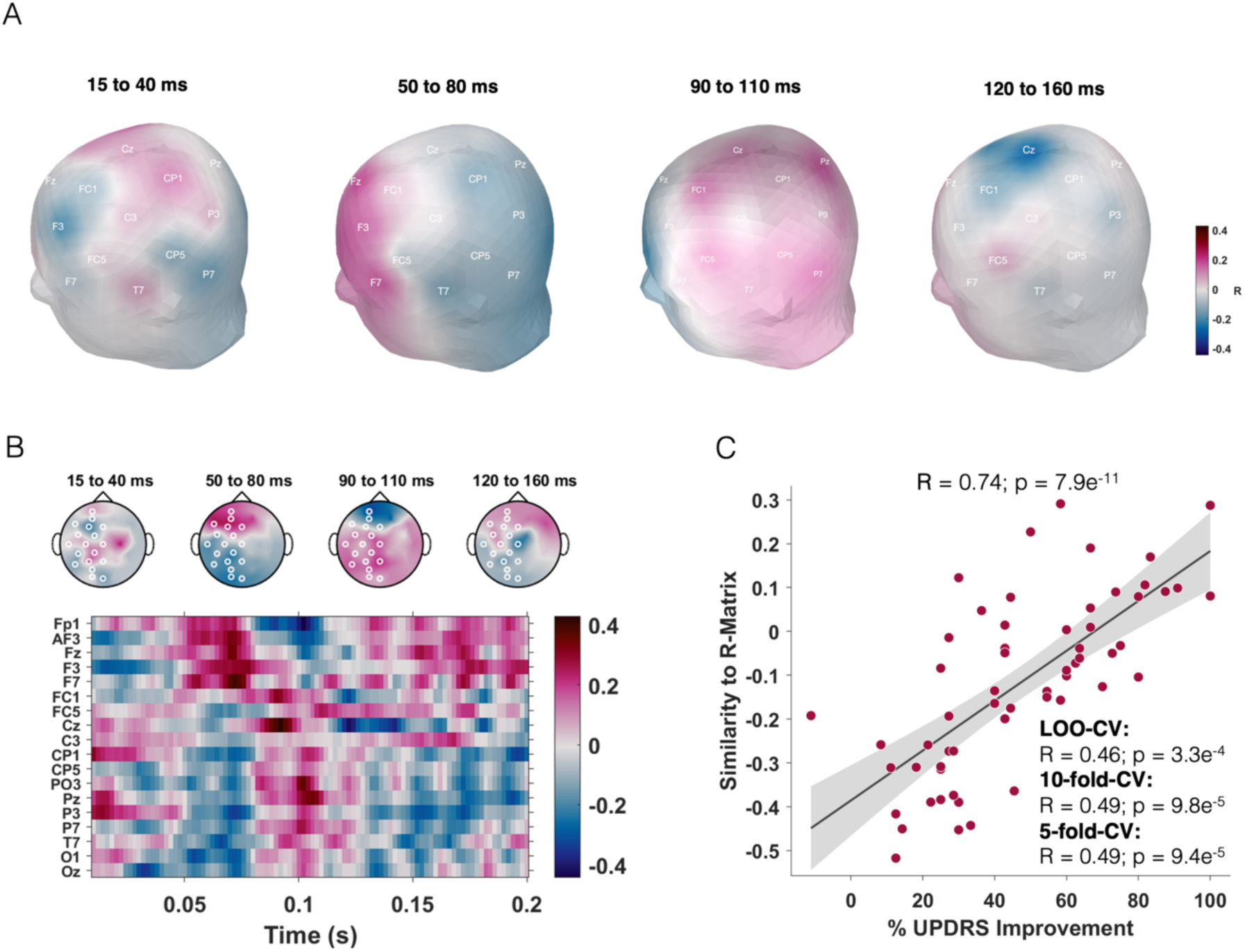
R-matrix and cross-validations. **A** Scalp topography of the correlation matrix (R-matrix) quantifying the relationships between evoked potentials (EP) and clinical improvements (N = 58 stimulated hemispheres) and averaged across the same time windows as previously defined. This topography approximates an ‘optimal’ stimulation response associated with maximal clinical improvements. **B** The same R-matrix across channels and time and respective 2D channel topographies. **C** Similarity estimates between each EP-map and the R-Matrix significantly correlated with empirical upper limb UPDRS-III improvement across the cohort in circular analysis and when subjected to various cross-validation designs. Grey shaded areas along the regression line represent the 95% confidence interval.

### DBS-evoked potential topography explains variance in stimulation outcomes

When correlating each EP-map with the R-matrix, the model explained ∼52% (R = 0.74; p = 7.9e^-11^) of variance within the whole sample. This analysis was circular and expresses the degree-of fit between data and model, reflecting the ceiling of how much variance the R-matrix could potentially explain. When subjecting the process to K-fold cross-validations (see Methods), the relationship remained significant (leave-one-hemisphere out design: R = 0.46, p = 3.3e^-4^; leave-one-patient out design: R = 0.41, p = 1.1e^-4^; 10-fold cross-validation: R = 0.49, p = 9.8e^-5^; 5-fold cross-validation: R = 0.49, p = 9.4e^-5^; see supplementary figure S3). This main analysis only considered ipsilateral and midline channels since the stimulation is known to produce EPs lateralized to the same hemisphere as the stimulated DBS electrode ^26,27^. However, when repeating the analysis including all 32 channels, cross-validations remained significant with slightly worse correlation coefficients (leave-one-hemisphere out design: R = 0.36 at p = 0.006; leave-one-patient out design: R = 0.30, p = 0.023; 10-fold cross-validation: R = 0.35, p = 0.007; 5-fold cross-validation: R = 0.37, p = 0.004; see supplementary figure S4).

### Segregated DBS-evoked potential topography for symptom-specific outcomes

In an exploratory analysis, we aimed at testing whether the topography of EPs could also shed light on *symptom-specific* outcomes. To do so, we repeated the main analysis substituting global motor improvements with improvements in three symptom domains; namely upper limb bradykinesia, tremor, and rigidity. This revealed three distinct R-matrices with differing topographies (Fig. 4). The rigidity R-matrix showed dominance in positively correlated frontal channels, while the one for tremor displayed a positive correlation of parieto-central channels. The bradykinesia R-matrix revealed a similar pattern to the rigidity R-matrix, while several more central channels with positive correlation coefficients emerged. Overall, unsurprisingly, the bradykinesia R-matrix resembled the one for global UPDRS-III improvements most. The general gradient observed across the three symptom domains, which orders symptoms as tremor → bradykinesia → rigidity from caudal to frontal, aligns well with symptom-specific results published in multiple literature findings^9,28,29^ (an example from a recent study that used tractography ^9^ is shown on the left for direct comparison in figure 4). Each symptom-specific model accounted for variance in symptom sub-score improvements (Rigidity: 83%; Bradykinesia: 49%; Tremor: 76%), however, these results are circular. When subjecting models to cross-validations, only the results for rigidity and bradykinesia remained significant, and much less so than for the global motor improvement model (supplementary figure S5). When each of the symptom-specific R-matrices was used to estimate the respective other two symptom improvements, the models failed to do so, except for the bradykinesia R-matrix estimating rigidity outcomes (supplementary figure S6).

**Figure 4.**
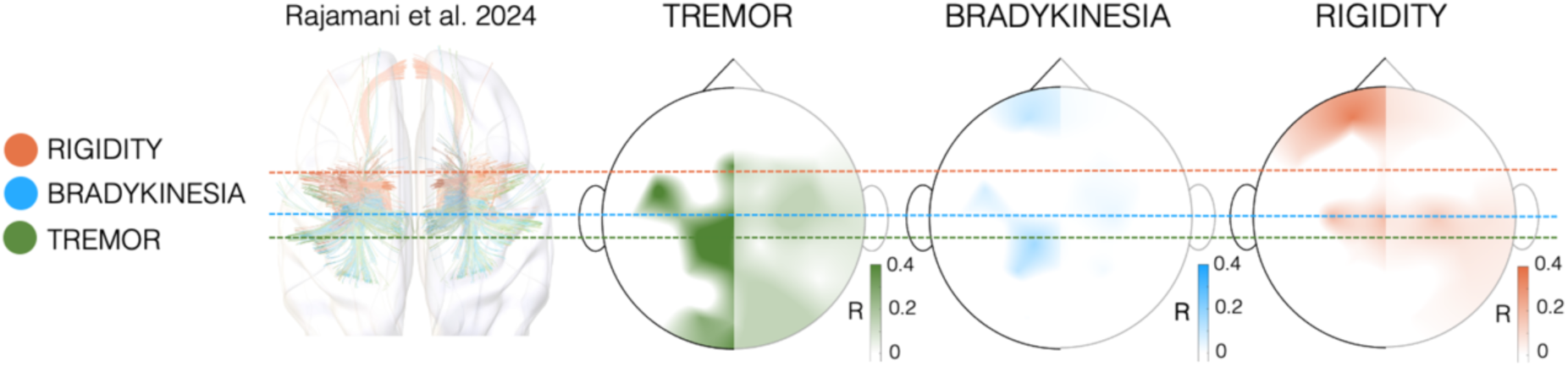
Symptom-specific correlation matrices (R-matrices) for the (dominant) N20 time point. The right side of the channel topography plot is blurred out (potentials are evoked by stimulation in the ipsilateral hemisphere, so all right-sided evoked potential recordings were flipped to the left before correlation, see Methods). The R-matrices show positive correlations only. The tremor R-matrix reveals positive correlations across parieto-central channels, while bradykinesia and rigidity display positive correlations at central and frontal channels. This topographical gradient aligns with the literature ^9,28,29^. An example of a recently published large-scale study that used tractography is shown in matching colors on the left ^9^. This study weighted fiber tracts from a normative structural connectome by symptom improvement. The horizontal dashed colored lines show the approximate weighted fiber peak for each of the three symptoms in relation to the R-matrix topography plots.

### DBS-evoked potential topography model outperforms imaging-based methods

Given the R-matrix method was able to explain variance in clinical outcomes robustly, we intended to compare it with other methods that have been used to estimate DBS outcomes, such as DBS electrode reconstruction models ^30^. To do so, DBS electrodes were localized (supplementary Fig. S1), and electric stimulation fields (e-fields) were estimated for all patients included within the discovery cohort based on clinically applied stimulation parameters using the default pipeline of Lead-DBS v3.0 ^30^ (see supplementary table 1 for stimulation settings). We then applied two methods to set models in relationship with clinical outcomes. First, Euclidean proximity between clinical stimulation contacts and the coordinate of a published motor improvement sweet spot within the STN were calculated ^31^. Additionally, weighted volume overlaps between e-fields with the STN were computed. These two metrics had shown correlations with clinical improvements in the past ^7,32^. As in these earlier reports, here, the two measures significantly correlated with clinical improvements (proximity to coordinate: R = 0.33; p = 0.012; volume overlaps with STN: R = 0.28; p = 0.036; see supplementary figure S7). However, in direct comparison with the EP-based approach, no significant amounts of additional variance in clinical outcomes were explained by either of the two imaging methods when combined in a linear model with EP-based estimates (EP-based approach as quantified by similarities to R-matrix from 10-fold-CV: β_std_ = 0.45; p = 3.9e^-4^; proximity to coordinate: β_std_ = 0.12; p = 0.346; volume overlaps: β_std_ = 0.20; p = 0.133; model R^2^ = 0.28; p = 6.2e^-4^).

### DBS-PULSE: DBS-evoked response pattern guides DBS contact selection

Finally, to explore the potential application of the proposed method to select optimal stimulation contacts (*DBS-PULSE* concept), we tested the R-matrix model in prospective dry EEG recordings. We recorded a more extensive dataset in three independent patients (5 hemispheres), acquiring EEG data for each of the four stimulation contact levels per hemisphere. This resulted in a total of 20 EP-maps which were again correlated with the R-matrix (generated based on the discovery cohort data, N = 58 hemispheres) to suggest optimal contacts. Frankly, each contact’s EP was spatially correlated with the R-matrix, and the resulting spatial correlation coefficients were used to rank the four contacts for each electrode. For each contact level, we determined the clinical effect threshold (minimum stimulation amplitude to reach a clinical improvement) as well as the therapeutic window (amplitude difference between clinical improvements and occurrence of side effects). When looking at each electrode separately, the method correctly ranked the DBS contact with the largest therapeutic window and lowest clinical threshold in all 5 hemispheres, which is significantly more than what can be expected by chance (p = 9.8e^-4^; binomial test under the null hypothesis that the algorithm selects a contact randomly with a 25% chance). Spatial correlation coefficients of each EP (test cohort) to the R-matrix model (discovery cohort) explained ∼44% of variance in clinical effect thresholds (R = 0.79; p = 0.002; note, that thresholds were inverted for ease of reading figures, see Fig. 5A), indicating, as expected, low effect thresholds for contacts with high similarity to the R-matrix. To account for repeated measures per hemisphere, we fitted a linear mixed-effect model with the similarity coefficients as fixed and hemisphere as random effect. This model explained significant amounts of variance in both clinical effect thresholds (β_std_ = 0.79; p = 1.16e^-4^) and therapeutic windows (β_std_ = 0.52; p = 0.03).

**Figure 5.**
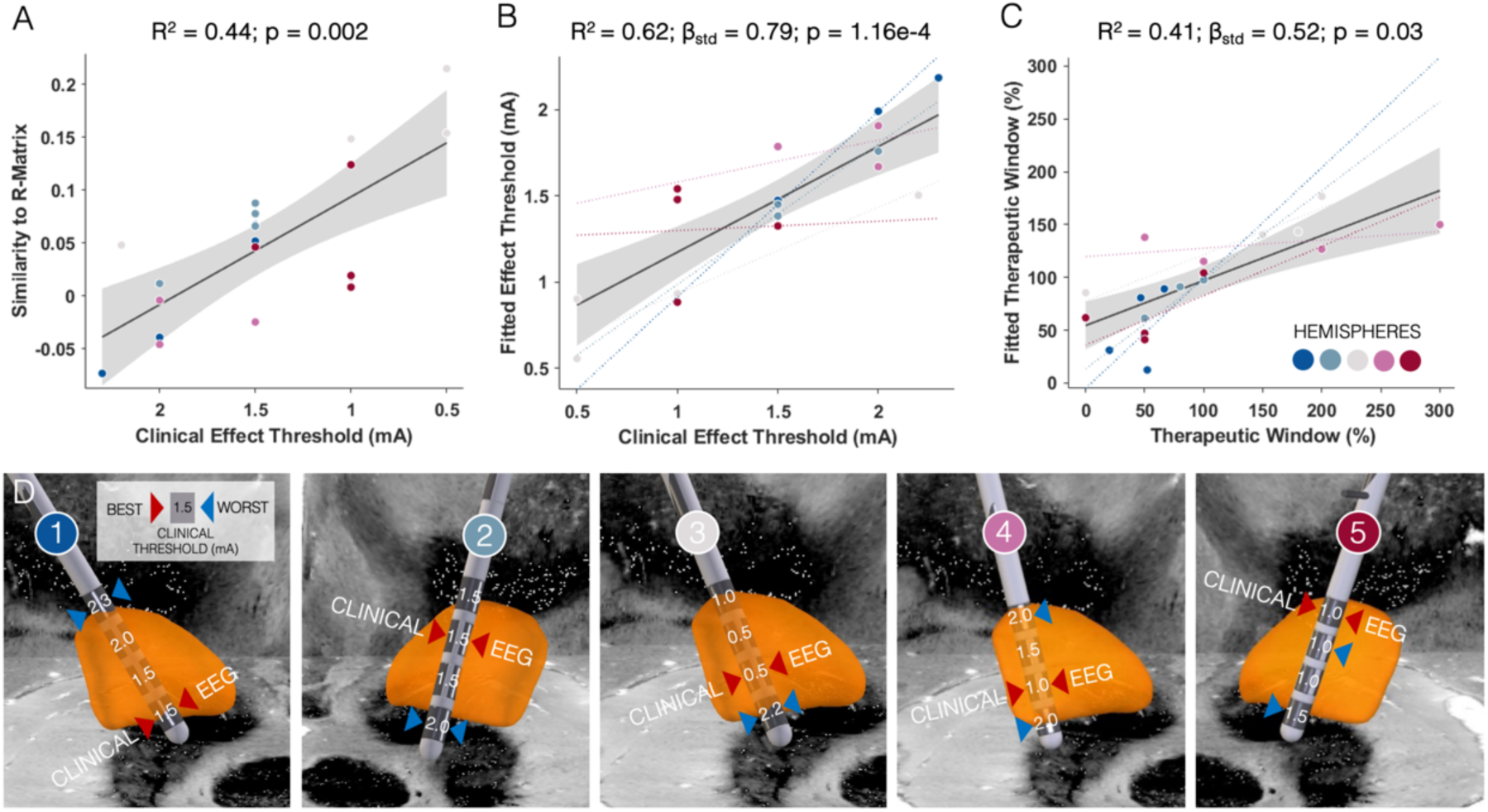
DBS-PULSE suggests optimal DBS contacts. **A** Correlation of clinical effect thresholds (minimum stimulation amplitude to reach clinical improvements) and similarity estimates between each DBS contact’s evoked potential (EP)-map and the correlation matrix (R-Matrix) from the N = 58 discovery cohort. The more similar the EP-map, the lower was the effect threshold for this particular deep brain stimulation (DBS) contact (x-axis inverted for ease of reading the figure). Dots are color-coded for each of the five hemispheres that were tested across three prospective patients. **B, C** Correlations of estimated (fitted) and empirical clinical effect thresholds and therapeutic windows (relative amplitude difference between clinical improvements and occurrence of side effects) when included in a linear mixed effect model that accounts for hemisphere as a random effect. The colored lines represent the fit within each individual hemisphere. **D** DBS electrode localizations labeled with the respective clinical effect thresholds. Arrows indicate the best (red), or worst (blue) clinical contact level(s) selected based on clinical testing (ground truth) and EEG-EP analyses (suggestion by the model).

To assess the method’s applicability in clinical practice, we wanted to evaluate whether recording durations as carried out here were necessary, or whether even shorter recording sessions could be equally informative. We therefore repeated the contact selection analysis for various recording durations (i.e. only using the first x seconds of recordings; see supplementary figure S8), ranging from less than 20 seconds (100 stimulation pulses/epochs) to about 2 and a half minutes (800 epochs). Besides the linear mixed effect model’s beta coefficients, we computed a receiver operating characteristic curve (ROC) for each recording duration. While beta coefficients remained stable for recording times of over 50 seconds (= 300 epochs), only at recording times of over 100 s (= 600 epochs), the ROC analysis began to indicate good discriminative abilities (AUC > 0.7), which is still significantly shorter than the recordings we carried out (∼960 epochs on average). Based on this analysis, recording times to suggest contact selections should at least have durations between 50 to 100 seconds per contact. This would amount to a total recording time between 6 and 10 minutes for 8 DBS electrode contacts.

## Discussion

Using a 32-channel dry EEG system, we recorded cortical potentials evoked by stimulation from 78 electrode contacts in 32 patients with Parkinson’s disease. The grand average EP revealed a fronto-central bipolar topography with peaks around 20, 60, 100, and 140 ms following the stimulation pulse. Based on our discovery cohort, which included 58 stimulation sites and a total of 57,503 stimulation epochs, we created a model of optimal EP topography (R-matrix). This optimal model approximates the EP pattern an effective electrode should ideally elicit to induce maximal symptom relief. Similarities of individual EP patterns to this optimal response model were able to estimate significant amounts of variance in empirical UPDRS-improvements, when subjecting the process to various cross-validation designs. When computing the R-matrices based on symptom sub-score improvements, we observed symptom-specific EP topographies, that were especially distinct for rigidity and tremor, while bradykinesia matched the global improvement model. In direct comparison to imaging-based measures, the EP-based model explained additional amounts of variance in empirical total UPDRS-III improvements. Finally, we tested whether the model was capable of selecting the optimal contact in prospective recordings where we activated each of four contact levels per electrode, independently. This process, which we term ‘PULSE-DBS’, correctly identified the best stimulation contact in five out of five tested hemispheres.

Several studies have investigated the mechanism of how EPs are generated. Potentials at latencies between 2 and 10 ms seem to result from an antidromic activation of the cortico-subthalamic ‘hyperdirect’ pathway which is also reflected by their cortical distribution ^6,33,34^. Potentials with longer latencies were hypothesized to instead originate from an orthodromic activation of the basal ganglia-thalamo-cortical loop and localized to the motor cortex and prefrontal areas ^6,34,35^. Importantly, the primary focus of our study was *not* to investigate mechanistic aspects of EP generation and was not geared towards analysis of early EPs (i.e. 2-10 ms). First, our recording setup did not allow an analysis of potentials with latencies shorter than 10 ms (supplementary Fig. 2 D), given artifacts from monopolar DBS shadow this time window. Additionally, short latency EP are only a few milliseconds long, while with our sampling rate of 500 Hz, only 2 samples were recorded per millisecond. Second, the spatial resolution of 32-channel dry EEG is limited and did not allow for a thorough differentiation of cortical sources underlying EP generation as previously studied ^6^. Third, to gain an actual mechanistic understanding of EP at the cortical level, subcortical alongside cortical recordings would be optimal ^36^.

Instead, the goal of our study was to test the *clinical utility* of a *setup* that could be *applicable in clinical practice*. We introduce a novel concept that utilizes the stimulation response pattern across the entire sensor space (and thus entire cortex) and is naïve to absolute magnitudes of EPs but rather leverages the relative distribution of EPs across the cortex as well as their time course. This being said, the data used in the algorithm was limited to longer latency EPs, namely as observed at 20, 60, 100 and 140 ms post-stimulus onset, while not restricting the analysis to any one chosen EP component.

Several studies before the present one have also focused on EP at latencies longer than 10 ms ^6,14,16,34,37^. These have been shown to correlate with clinical improvements along with short latency potentials in patients with Parkinson’s disease ^6,16,37^. Long-latency potentials have further been conceptualized to occur from resonant phenomena within the basal ganglia-cortical circuitry, with a preferential propagation of activity around ∼20 Hz ^26,37^. The frequency of the grand average EP that we recorded only roughly matched a 20 Hz resonant oscillation (Fig. 2). The R-matrix, however, showed a more pronounced peak around 100 ms across parietal and central channels alongside a frontal 60 ms and a central 20 ms peak, which would constitute a ∼25 Hz resonant oscillation.

Critically, besides the resonance of EP (time axis), our model was informed by the *topography* changes (spatial/channel axis) across time. Consistent with previous work ^6,26^, the N20 was distributed along fronto-central channels. The EP topography at longer latencies has only been studied with a few EEG channels so far, with the largest amplitudes recorded in frontal channels ^14^, which is also in agreement with our results (Fig. 2B). When correlating the EP-maps with clinical improvements, the pattern for the N20 changed toward a positively correlated band across central channels (Fig 3 A). Previous findings indicated relationships between N20 amplitudes and motor improvements for motor cortex and supplementary motor area but not for other frontal areas ^6^. Additionally, the R-matrix highlights a positively correlated pattern at parieto-central channels around 100 ms. In the grand average, there was no pronounced peak around this latency. This could point toward a clinically beneficial EP pattern, that is dominated by high beta resonance with an inter-peak distance of about 40 ms.

When computing symptom-specific R-matrices, we observed three distinct ‘optimal’ EP topographies that were dominated by frontal correlations in the case of rigidity and parieto-central correlations for tremor (Fig. 4). This symptom-specific gradient aligned with prior literature results, ^9,28,29^ including a recent large-scale study that investigated a total of 237 patients ^9^. Notably, the gradient observed for EEG recordings at channel and scalp level is not well resolved anatomically and can only be interpreted as a rough estimate of the actual cortical distribution. Still, our data suggest, that the segregated topography could help guide a more personalized programming approach that could go above and beyond optimizing global motor improvements. Such an approach could further be informed by baseline symptom sub-scores, yielding a personalized symptom-weighted model for individualized DBS parameter selection as previously suggested in the framework of ‘network blending’ ^9,38^.

Our study addressed several aspects of clinical applicability for EP-guided DBS programming. First, the 32-channel dry EEG system tested to record DBS-EP can be set up within a few minutes and appears robust against monopolar DBS pulses, allowing us to study EP as early as 20 ms after the stimulation pulse (supplementary Fig. 2 D). Second, even though medication effects on longer latency potentials have been described ^26^, we purposefully chose to record patients in the condition in which they would usually visit an outpatient department (best medication ON) to potentially prevent the necessity of a medication off-state for an EP-guided programming approach. Third, we tested the downscaling of the recording time to enable a relatively quick EP recording during or shortly before an outpatient clinic appointment. Hypothetically, with a fixed stimulation amplitude for all contacts, the process of contact selection could be concluded within about 10 minutes (50-100 seconds per contact) for 8 DBS electrode contacts based on our recording duration analysis (see supplementary figure S8) after a ∼2-5 minute set up time. Fourth, the analysis pipeline was kept simple to enable an easy and automated approach to data cleaning and results. Even when considering the time for manual EEG data analysis steps, with 10 minutes of EEG data, the amount of time for clinical programming and testing with bothersome symptoms and side effects for the patient would still be remarkably reduced. Additionally, the model seems to be generalizable toward novel data, as shown using both cross-validations and hold-out test set application. In other words, based on these results, no further training of the model would be necessary to apply it to unseen cases.

Despite these advantages of the approach, several limitations should be considered when interpreting results. First, we only focused on upper limb UPDRS improvements when evaluating patients alongside the dry EEG recordings. This might reduce the applicability of our results to lower limb motor function and axial symptoms such as gait. Still, in many centers, upper limb symptoms are exactly what is being measured during monopolar review testing, i.e. the standard of care in the first stage of programming does not take other symptoms into account, either. Second, our approach will be limited to contact selection and given the fixed amplitude and frequency design of the experiment, no inferences on the optimal stimulation amplitude or frequency can currently be made. Yet, even with contact selection alone, the approach would present a useful clinical tool to speed up DBS programming. Moreover, in disorders in which stimulation effects are less immediate or even prolonged, such as dystonia or depression, biomarkers for contact selection and therapeutic success could considerably improve the success of DBS ^39–41^. Still, further studies are needed to test the applicability of our approach in other disorders and symptoms above and beyond the motor domain, and different R-matrix models need to be trained for other disorders and DBS targets. Third, our prospective results are limited to 20 DBS contacts sampled in only five hemispheres (three patients). Future studies to prospectively validate the approach in a double-blind randomized clinical trial will be necessary to assess non-inferiority or even superiority to standard of care clinical testing using monopolar reviews. Fourth, even though our method selected the correct clinical contact level in all of the five hemispheres, we did not test for directional DBS contacts and most of the patients in the discovery cohort were stimulated with omnidirectional stimulation settings. Previous work suggests that DBS-EP might be capable of indicating the optimal stimulation direction in segmented leads ^16^. However, differences across segmented contacts were small and future studies with a model trained on both directional and omnidirectional contacts are needed. Lastly, our approach could further benefit from advanced signal processing methods, such as spatial filtering, that could enhance the model’s performance and enable detecting e.g. small differences of spatial patterns between directional DBS contacts ^42^.

In conclusion, we present a novel approach, *DBS-PULSE*, that proved useful for identifying the optimal stimulation contact and would be clinically applicable at scale. Future validations in larger patient populations will be critical to further confirm findings on our model’s performance for DBS contact selection.

## Methods

### Recruitment and clinical data collection

We recruited 32 patients (11 female; mean age: 60.4 ± 6.8 years) with Parkinson’s disease from the Department of Neurology at Charité – Universitätsmedizin in Berlin, Germany and obtained written informed consent of all patients prior to study inclusion. Three out of 32 patients were recruited upon completing the primary analysis and recruitment of the discovery cohort for prospective validation (test cohort) and underwent more extensive recordings (see below). The study protocol is in accordance with the Declaration of Helsinki and was approved by the local ethics committee at Charité (Ethic proposal number EA2/247/21). LEDD were calculated based on the standard procedure described in the literature ^43^ and individual LEDD, chronic DBS settings and other patient demographics are reported in supplementary table S1.

### Data acquisition

We used a 32-channel dry EEG system (Starstim 32, Neuroelectrics, Cambridge, MA, USA) and acquired resting state data referenced to the right ear electrode with a sampling rate of 500 Hz. In a discovery cohort of 29 patients, we recorded dry EEG data during unilateral STN stimulation at the individual clinically established stimulation contact (see supplementary table S1) with a frequency of 2 Hz and a stimulation amplitude of 8 mA for 10 minutes in each hemisphere (N=58). Given the low stimulation frequency, these amplitudes were tolerated well by the participants. We purposefully chose high stimulation amplitudes to elicit strong and sustained EP over the time course of the inter-pulse interval. During stimulation of the left STN, the right STN electrode was turned off and vice versa. In the discovery cohort, the stimulation frequency was set to 2 Hz to enable the analysis of long-latency EP ^37^. This decision was also related to the fact that we kept the clinically used monopolar stimulation montage at the chronic stimulation contact and that together with the relatively low sampling rate these artifacts might prevent the analysis of short latency EP ^6,13,34^. After the recording, UPDRS upper limb motor scores were assessed (UPDRS-III items 3.3-3.6; 3.15-3.17) under stimulation switched off and after at least 30 minutes of high-frequency stimulation at chronic DBS settings. To characterize potential artifacts affecting the dry EEG recordings, we produced a conductive gelatine phantom for combined EEG-DBS recordings. An openly available phantom model for EEG recordings was used to print a mold using a commercially available 3D printer ^44^. Two DBS electrodes were introduced into the gelatine and connected to an impulse generator (supplementary Fig. S2 B). Stimulation was applied with the same settings as in the patient recordings.

In three of the 32 recruited patients, that were not included in the discovery cohort, we prospectively recorded a validation dataset. To reduce the recording time, we increased the stimulation frequency to 6 Hz and recorded 2.5 minutes of dry EEG data per stimulation contact level (10 minutes per hemisphere). This adjustment was justified by our observation in the discovery cohort that no clear EP was detected after 200 ms. In one of the three patients, we only acquired data in one hemisphere due to extended recording time caused by reference electrode issues, which were resolved but which prolonged the session. Consequently, we could complete data acquisition for only one hemisphere before the patient became too tired to continue without the clinically effective stimulation. We determined the clinical effect thresholds for each electrode level in the three patients based on the earliest clinically relevant improvement of contralateral upper limb bradykinesia and rigidity as well as side effect thresholds at the earliest sustained side effect in a separate testing session in medication off-state (standard of care).

### Data analysis

The EEG data of patient and phantom recordings was analyzed with Brainstorm ^45^. The data was visually inspected for artifacts and bad segments and channels were rejected. Subsequently, bad channels were interpolated using the surrounding channels. We applied a high pass filter at 4 Hz. This relatively high filter frequency was chosen, given the time scale of our EP analysis with a time range of interest of 100 to 200 ms, which is considerably shorter than the temporal dynamics captured by these low frequencies. We then re-referenced the data to the average across channels. We used the custom peak detection algorithm as implemented in Brainstorm to detect the stimulation artifact with a threshold of 2 standard deviations and a minimum inter-stimulus interval of 490 ms (150ms for the 6 Hz DBS recordings) in a visually selected channel, that showed the largest stimulation artifact. Then, we epoched the data based on the detected stimulation artifacts with a baseline period of 100 ms before the stimulation pulse and 400 ms of data after (16 ms before and 150 ms after in the case of 6 Hz DBS in the test cohort data). After the rejection of bad segments (20.7 % of epochs) an average of 958 (± 236) epochs were included (total number of epochs across all recordings: 57,503). The average across epochs was calculated per hemisphere resulting in a total of 58 amplitude maps (EP-maps): 32 channels x time [-50, 450 ms] (see Fig. 1 A). The phantom EEG data was processed using the same pipeline.

To compare the stimulation artifact between phantom and patient recordings, the absolute EP were averaged across all EEG channels for the phantom and two representative patient recordings (supplementary Fig. 2). Then, a logarithmic scale was chosen to show the stimulation artifact alongside the EP on one scale (supplementary Fig. 2). Given the slightly different topography of the DBS artifact in phantom and patient recordings, it was necessary to average across channels and to use absolute amplitudes to be able to make artifacts comparable. We additionally generated an EP map for both phantom and patient recordings and overlaid the two to visualize artifact and EP across different channels. One of the two representative patients was selected and the average across epochs for the right STN stimulation was used to create an EP-map (Fig. 1 A). Then, the same steps were applied to a right sided phantom stimulation file. We calculated a mask across time points and channels indicating where the phantom EP-map showed a higher amplitude than the patient recording for the respective channel and time point. Then we applied the mask to the phantom file, by setting the respective values to 1 and every other value to 0. This mask was then plotted on top of the patient EP-map (Fig. 2 E).

The 58 average EP-maps were now used to create an ‘optimal’ EP-map, indicating the pattern and amplitude across channels and time that is associated with UPDRS improvement (Fig. 1B). This ‘optimal’ map was created using an element wise correlation with the percentage improvement in contralateral hemi-body UPDRS scores following a mass-univariate model approach ^23^. Before correlating the EP-maps with clinical improvement, we masked them to the relevant time window between 10 ms and 200 ms after the stimulation pulse (10 ms and 150 ms for the prospective recordings with 6 Hz DBS) and flipped the maps by assigning right to left channel labels for the right STN-DBS recordings (e.g. channel signal C4 was assigned to C3 etc.). After all right stimulation recordings were flipped to the left, we selected the left-sided channels (Fp1, AF3, C3 etc.) as well as the midline channels (Fz, Cz, Pz and Oz) for further analysis (= 18 channels). These steps were also performed for the prospective patient recordings. An additional analysis with all 32 channels is shown in supplementary figure S4.

We repeated the element-wise correlation with the percentage improvement in contralateral upper limb symptom sub-scores (rigidity: 3.3 b-c; bradykinesia: 3.4-3.6 and tremor: 3.15-3.17) and plotted the R-matrices (Figure 4 and supplementary figure S5) for exploratory analysis. For each of the symptom improvements, patients with baseline symptom scores of less than 2 points were excluded. Hence, for rigidity a subset of 21 and for tremor a subset of 8 hemispheres were included. As the defining symptom for Parkinson’s disease, bradykinesia was present in all patients at baseline (N=58).

### DBS electrode localization and imaging-based analysis

DBS leads were localized using Lead-DBS v3.0 for all patients as previously described ^30,46^. Postoperative CT and preoperative MRI were co-registered and non-linearly warped to ICBM 2009b Nonlinear Asymmetric (‘MNI’) space using Advanced Normalization Tools (ANTs, https://stnava.github.io/ANTs/). Following brain shift correction due to possible pneumocephalus, the non-linear warp was manually refined with focus on the STN region as the target zone of interest using WarpDrive ^32,47^. Then, electrode trajectories were reconstructed using PaCER ^48^ in case of postoperative CT images or TRAC/CORE ^49^ if postoperative MRI scans were available. Last, the chronic DBS settings (supplementary table S1) were used to compute the Euclidean proximity to published motor symptom improvement sweet spot coordinates in MNI-space ([± 12.42, -12.58, - 5.92 mm]) ^31^. Additionally, we computed the volume overlap of electric stimulation fields (E-fields) with the STN in native space. To this end, the chronic DBS settings were used to compute E-fields using recently developed methods and the overlap with the STN outline in native patient space was computed for each hemisphere and correlated with % UPDRS improvements ^50^.

### Statistics & model Validations

The correlation matrix (R-matrix) has the same dimensions as the masked EP-map and consists of correlation coefficients indicating the relationship to motor outcomes across hemispheres (18 channels x time [10, 200 ms]). To test how much variance the R-matrix is able to explain in this dataset, we correlated all of the individual EP-maps with the R-matrix based on the full cohort. The resulting correlation coefficients (i.e. spatial correlations/similarities) can serve as estimates of the empirical improvements. We thus correlated empirical improvements and similarities to the R-matrix to calculate the ceiling of variance explained by the model. Since we use the same EP-maps to generate the R-matrix that we later correlate it with, this analysis is circular and can solely express the overall variance that can be explained by the model. Thus, to validate the R-matrix model, we subjected it to leave-one hemisphere-out cross-validation. The similarity between each left out hemisphere’s EP-map and the R-matrix was calculated as an estimator of clinical improvement by correlating the two matrices. Iteratively, the process was repeated, each time leaving out one hemisphere from the R-matrix calculation. The similarities to the respective R-matrices were then correlated with the empirical improvements across all hemispheres. To account for potential similarities between the two EP-maps based on left and right sided stimulation recorded in the same patient, we performed a leave-one-patient out analysis as well. Additionally, the data was subjected to a 10-fold and 5-fold cross-validation essentially shuffling the data to 10 or 5 folds, building an R-matrix with all but one-fold and using the latter to calculate the similarities to the R-matrix (see supplementary figure S3 and S4). All analyses were carried out using a two tailed correlation test (Spearman’s rho).

Also, the symptom-specific R-matrices were subjected to cross-validations. For the tremor R-matrix, only leave-one-out cross-validations could be applied, given the small sample size. Given the hypothesis-driven, exploratory nature of the symptom-specific R-matrix analysis and given that in cross-validation experiments, only positive correlations are valid, we employed a one-sided (right-tailed) significance test for the correlation. Additionally, the symptom-specific R-matrices were used to estimate improvements in each of the other symptoms. To this end, each symptom R-matrix was correlated with each individual EP-map in a leave-one-out design and the three similarity values per hemisphere were each correlated with tremor, rigidity and bradykinesia outcomes resulting in a 3 x 3 matrix (see supplementary figure S6).

To compare the R-matrix approach with two imaging-based methods to estimate DBS outcomes, we computed the Euclidean proximity between clinical stimulation contacts and the coordinate of a published motor improvement sweet spot within the STN as well as the weighted volume overlaps between e-fields with the STN (see above). These two metrics were then correlated with the percentage UPDRS-III improvements using two tailed correlation tests (Spearman’s rho). To directly compare the amounts of variance in DBS outcomes that were explained by each of the imaging-based and the EEG method, we fitted a linear model. In this model, we included (1) the spatial correlations of individual unseen EP-maps with the R-matrix from the 10-fold-cross validation (see above), (2) the Euclidean proximities, as well as (3) the weighted volume overlaps as independent variables and the percentage UPDRS-III improvement as the dependent variable.

To test if the R-matrix can be used to determine the optimal stimulation contact in unseen patients based on dry EEG recordings (DBS-PULSE concept), we used the prospective validation dataset consisting of EP-maps for 20 contacts in five hemispheres. We correlated the 20 EP-maps with the R-matrix that was based on the discovery cohort to estimate improvements for each of the contacts. We then correlated the similarities to the R-matrix with therapeutic effect thresholds previously determined for each of the contacts (Fig. 5 A). To account for the repeated recordings per hemisphere, we additionally fitted a linear mixed-effect model to relate R-matrix similarities (fixed effect) to therapeutic thresholds in mA and relative therapeutic windows in % (therapeutic window in mA divided by therapeutic threshold in mA). Hemisphere was modelled as a random effect. We determined the optimal contact based on the lowest clinical threshold and the largest therapeutic window and compared that contact to the DBS contact that showed the highest EP-map similarity to the R-matrix. To assess whether this selection (based on the EP-map’s similarity) was better than chance, we conducted a binomial test under the null hypothesis that the algorithm selects a solution randomly (p = ¼= 25%; i.e. 1 out of 4 contacts), and estimated the probability of obtaining five correct selections out of five trials by chance, which we report as a p-value.

To determine if the time for dry EEG recordings could be reduced, we repeated the linear mixed effect model analysis for different numbers of epochs from the full dataset (100, 200, 300, 400, 500, 600, 700, and 800 epochs), equivalent to recording durations of ∼20 to 130 seconds, when a stimulation frequency of 6 Hz was applied. Additionally, we conducted an exploratory receiver operating characteristic-(ROC) analysis to assess the performance of our model at each of the recording durations. This analysis was conducted based on a contact selection variable (considering both, lowest clinical threshold and largest therapeutic window) and used the similarity of the respective contact’s EP-map for contact selection with the highest similarity indicating the best DBS contact.

## Data Availability

All data produced in the present study are available upon reasonable request to the authors

## Acknowledgements

This work was supported by the Deutsches Zentrum fur Luft- und Raumfahrt (DynaSti grant within the EU Joint Programme Neurodegenerative Disease Research, JPND). BHB and LLG gratefully acknowledge support by the Prof. Dr. Klaus Thiemann Foundation (Parkinson Fellowship 2022 and 2023). A.H. was supported by National Institutes of Health (R01MH130666, 1R01NS127892-01, 2R01 MH113929 & UM1NS132358) as well as the New Venture Fund (FFOR Seed Grant). GW acknowledges support by Deutsche Forschungsgemeinschaft (DFG) Project number 424778381 (TRR 295) and Project number 511192033. EF received funding from the Deutsche Forschungsgemeinschaft (CRC 295, FL 760/6-1) and Volkswagen foundation (Lichtenberg program).

## Supplementary Material

**Supplementary Table 1.** Patient demographics. Sex, Age in years, LEDD in mg, DBS parameters for left and right STN (STN L and STN R), UPDRS III items 3.3-3.6; 3.15-3.17 scores for Stimulation Off and On state, DBS system (MDT=Medtronic, BSC= Boston Scientific). Mean and standard deviation are provided for Age, LEDD and UPDRS scores. LEDD (levodopa equivalent daily dose) in mg was calculated based on Jost et al. Mov Dis 2023.

|  |  | UPDRS |  |  | DBS Parameters |  |  |
| --- | --- | --- | --- | --- | --- | --- | --- |
| ID | Sex | Off | On | LEDD | STN L | STN R | Electrode model |
| 01 | f | 15 | 11 | 390 | 3- / C+; 1.4 V | 1- / 0,2+; 1.3 V | MDT 3389 |
| 02 | m | 15 | 12 | 665 | 4,5- / C+; 3.0 mA | 4,5- / C+; 3.1 mA | BSC Vercise |
| 03 | m | 19 | 13 | 739 | 1abc- / 2abc+; 3.6 mA | 1abc-/2abc+; 3.6 mA | MDT B33005 |
| 04 | m | 21 | 8 | 100 | 1abc,3- / C+; 1.0 mA | 3abc- / C+; 1.6 mA | MDT B33005 |
| 05 | m | 17 | 17 | 675 | 1abc- / C+; 1.5 mA | 1abc- / C+; 2.6 mA | MDT B33005 |
| 06 | m | 22 | 9 | 450 | 2abc- / C+; 1.0 mA | 2ab- / C+; 3.6 mA | MDT B33005 |
| 07 | m | 23 | 14 | 475 | 2- / C+; 1.3 mA | 2,3,4- / C+; 3.6 mA | BSC Vercise |
| 08 | m | 18 | 6 | 526 | 2,3- / C+; 2.6 mA | 3,4- / C+; 2.6 mA | MDT 3389 |
| 09 | f | 14 | 9 | 839 | 2abc- / C+; 1.4 mA | 1- / C+; 3.5 mA | MDT B33005 |
| 10 | m | 21 | 15 | 575 | 3abc- / C+; 1.9 mA | 2abc- / C+; 1.9 mA | MDT B33005 |
| 11 | m | 11 | 5 | 805 | 7,8,9- / 4,5,6+; 2.7 mA | 7,8,9- / 4,5,6+ / 2.2 mA | BSC Cartesia |
| 12 | f | 18 | 15 | 495 | 2 to 7- / C+; 3.9 mA | 2,3,4,5,6,7- / C+; 1.8 mA | BSC Vercise |
| 13 | m | 23 | 5 | 980 | 1,2abc- / C+; 2.3 mA | 1abc,2abc- / C+; 2.3 mA | MDT B33005 |
| 14 | f | 11 | 2 | 600 | 2abc- / 3+; 3.0 mA | 1abc- / C+; 2.5 mA | MDT B33005 |
| 15 | m | 26 | 22 | 575 | 5,6,7- / C+; 3.0 mA | 5,6,7- / C+; 2.0 mA | BSC Vercise |
| 16 | m | 21 | 13 | 1114 | 1- / C+; 4.4 mA | 3- / C+; 3.8 mA | MDT 3389 |
| 17 | f | 22 | 15 | 235 | 2c-,3- / C+ 2.4 mA | 1a,2a,3- / C+; 2.4 mA | MDT B33005 |
| 18 | m | 15 | 6 | 750 | 1abc- / C+; 1.5 mA | 1abc- / C+; 1.5 mA | MDT B33005 |
| 19 | f | 25 | 16 | 352 | 2abc,3- / C+; 3.2 mA | 2abc / C+; 2.4 mA | MDT B33005 |
| 20 | m | 22 | 8 | 325 | 0- / C+; 3.1 mA | 1- / C+; 2.2 mA | MDT 3389 |
| 21 | m | 18 | 8 | 300 | 1abc- / C+; 2.2 mA | 1abc- / C+; 1.4 mA | MDT B33005 |
| 22 | m | 23 | 5 | 725 | 1abc- / C+; 2.4 mA | 1abc- / C+; 2.4 mA | MDT B33005 |
| 23 | m | 10 | 3 | 400 | 2abc- / C+; 1.5 mA | 2abc- / C+; 1.5 mA | MDT B33005 |
| 24 | f | 27 | 6 | 1000 | 1abc- / C+; 2.0 mA | 1abc- / C+; 2.4 mA | MDT B33005 |
| 25 | f | 21 | 9 | 937 | 1- / 2+; 2.9 mA | 2- / 1+; 1.9 mA | MDT 3389 |
| 26 | m | 16 | 10 | 1075 | 0- / C+; 1.4 mA | 2abc- / C+; 1.8 mA | MDT B33005 |
| 27 | m | 19 | 13 | 350 | 2abc- / C+; 2.5 mA | 2abc- / C+; 3.4 mA | MDT B33005 |
| 28 | f | 16 | 10 | 525 | 1abc- / C+; 2.5 mA | 1abc- / C+; 2.8 mA | MDT B33005 |
| 29 | f | 20 | 5 | 200 | 1abc- / C+; 2.5 mA | 1abc- / C+; 2.9 mA | MDT B33005 |
| 30 | m | - | - | 1075 | 1abc- / C+; 1.9 mA | 2abc- / C+; 1.7 mA | MDT B33005 |
| 31 | f | - | - | 525 | 1abc- / C+; 1.5 mA | 1abc- / C+; 1.5 mA | MDT B33005 |
| 32 | m | - | - | 798 | 1a-,2a- / C+; 3.6 mA | 1a-,2a- / C+; 4.5 mA | MDT B33005 |
| **Mean** | **11 f** | **18.9** | **10.0** | **612** |  |  |  |
| **Std** |  | **4.4** | **4.7** | **273** |  |  |  |

**Supplementary Figure S1.**
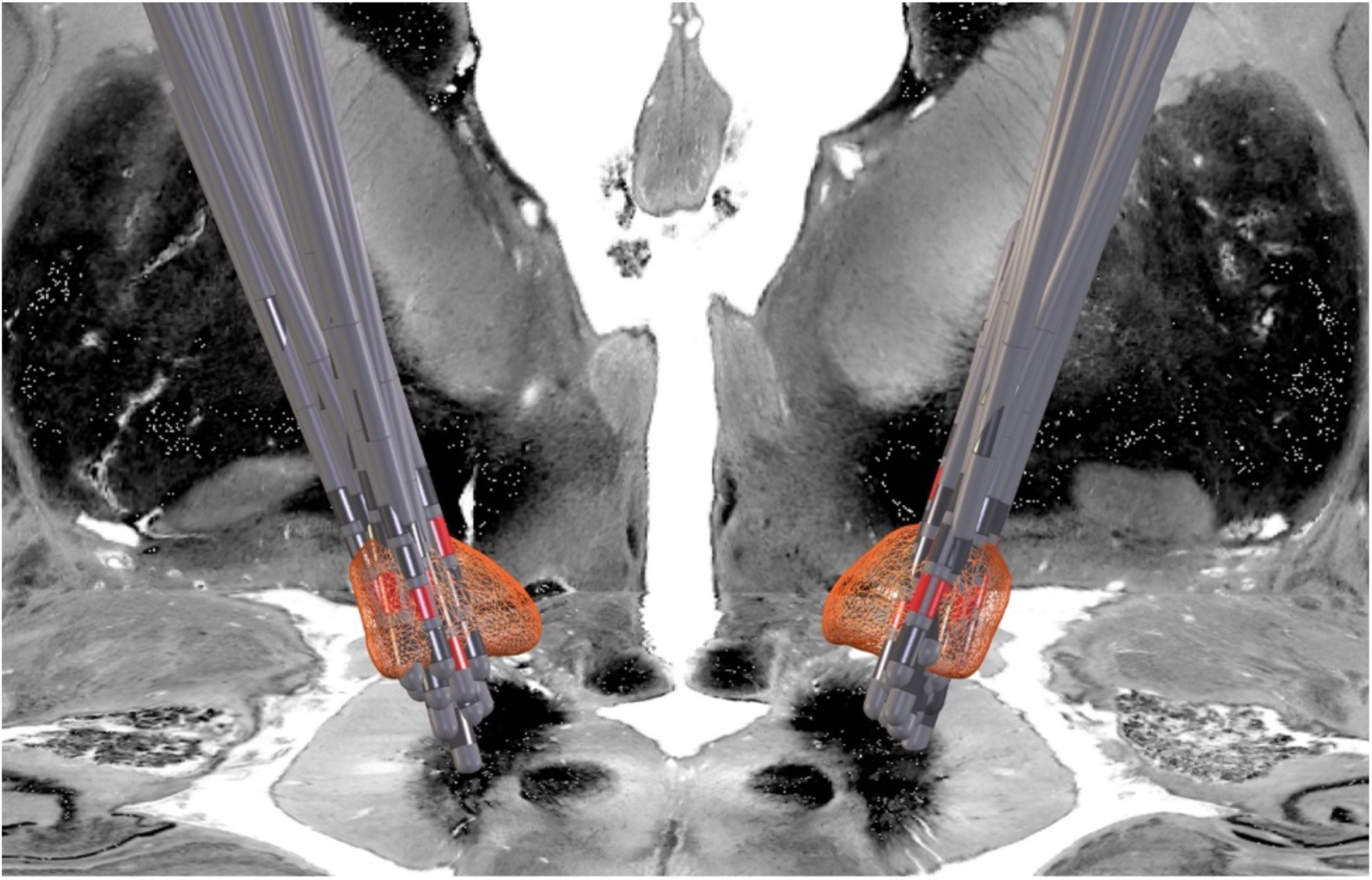
DBS electrode localization (view from posterior-superior). DBS electrodes (N = 32) are visualized in MNI-space within both STN and chronic stimulation contacts are highlighted in red. As a backdrop image the Big Brain Atlas was used (Amunts et al. 2013).

**Supplementary Figure S2.**
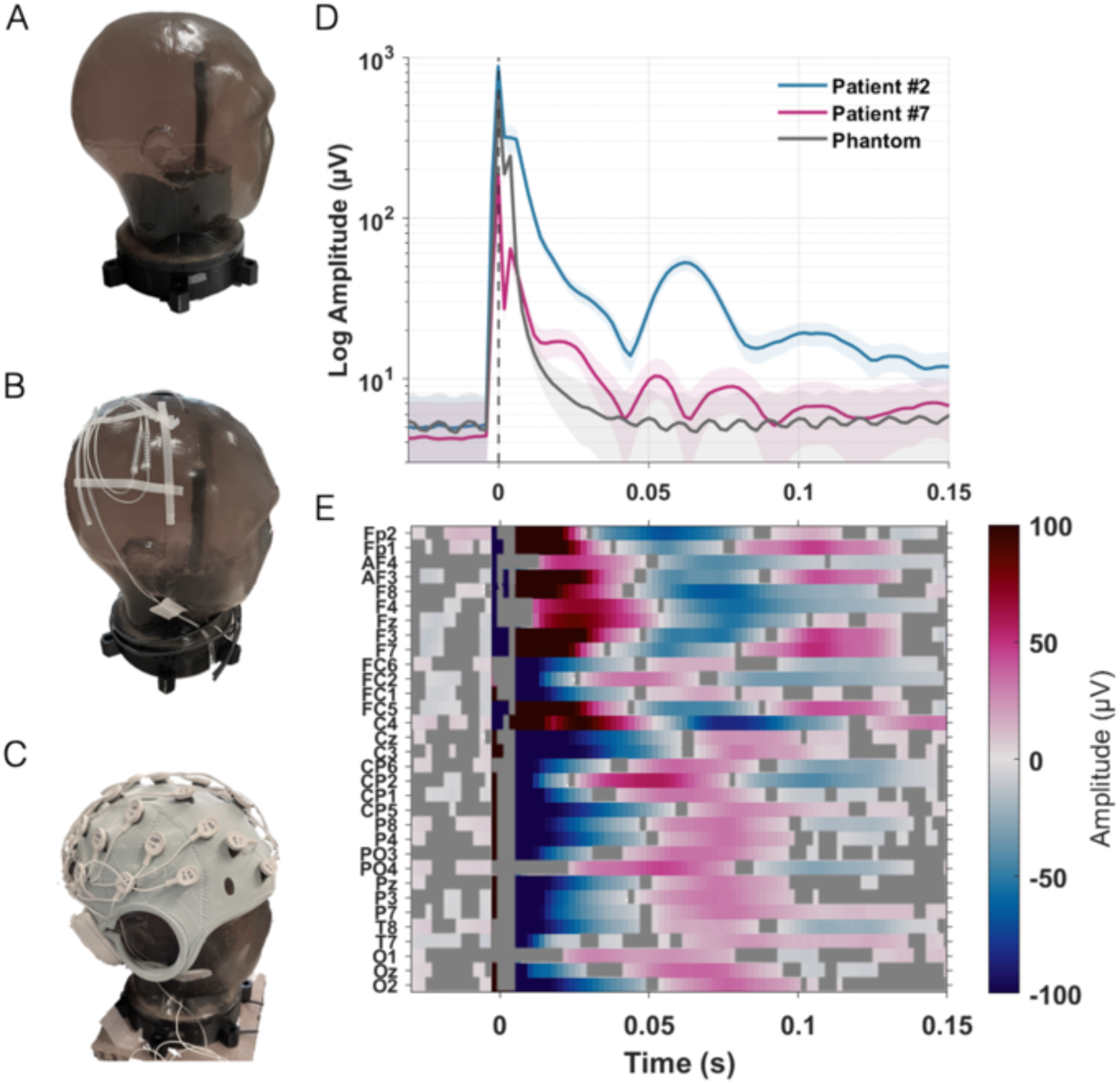
A-C: Dry EEG phantom setup. A: gelatine EEG phantom produced using a 3D-printed mold ^44^. B: Deep brain stimulation (DBS) electrodes introduced in gelatine and extension cables attached to the phantom head. C: 32-channel dry EEG cap on the conductive gelatine phantom. D: DBS artifact comparison between phantom recording (grey) and two representative patient recordings (see below). In the graph the average across channels and the standard error of the mean (shaded areas) are depicted. The amplitude is displayed on a logarithmic scale to visualize the high amplitude of the stimulation artifact alongside the evoked potential (EP). The 20 ms – response (N20) may be distinguished from the artifact decay after around 10 ms following the DBS pulse (time point 0). E: Average DBS-EP in a representative patient across dry EEG channels and time with color-coded amplitudes. The grey overlay represents the time periods in which the phantom recording had a higher amplitude than the patient recording for the respective channel and time point.

**Supplementary Figure S3.**
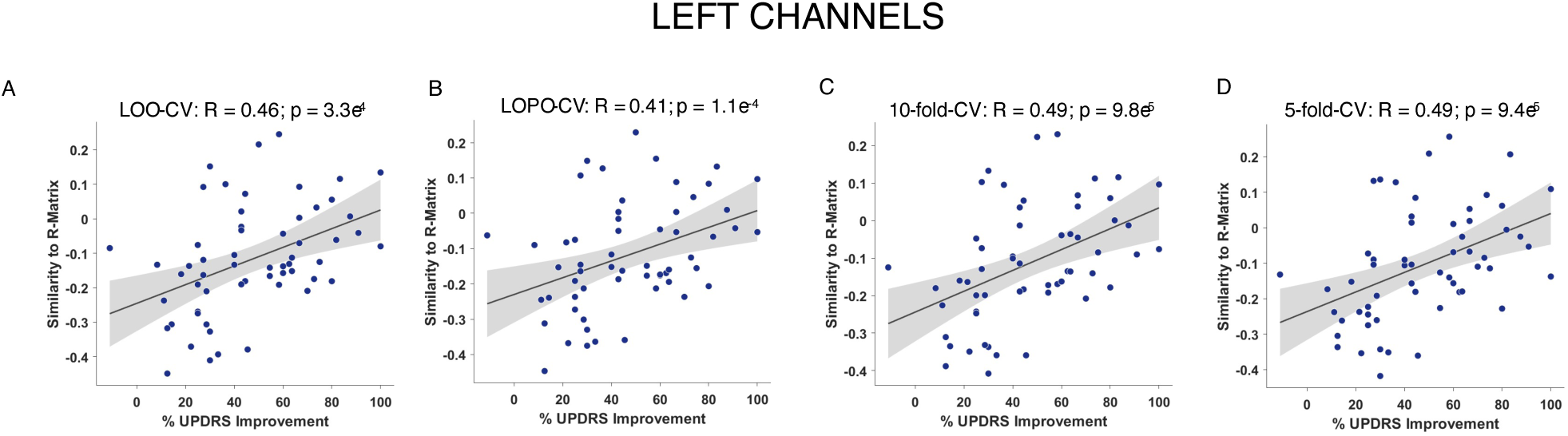
Cross-validation results using left channels. Correlation plots for each of the cross-validations using similarities to the R-matrix. LOO-CV= leave-one-hemisphere out cross-validation; LOPO-CV= leave-one-patient out cross-validation.

**Supplementary Figure S4.**
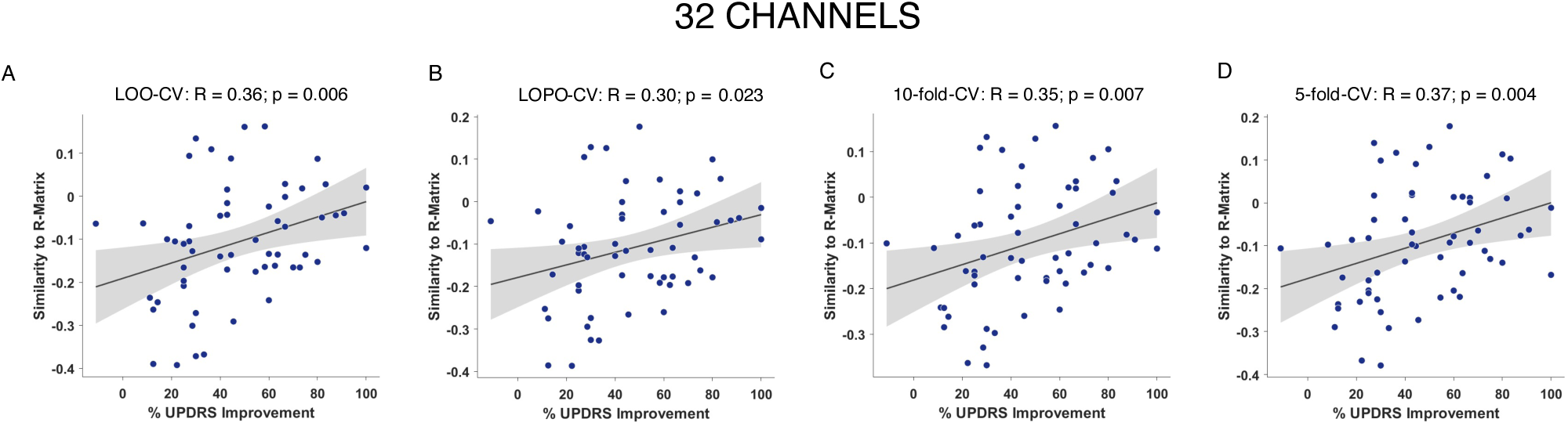
Cross-validation results using all 32 channels. Correlation plots for each of the cross-validations using similarities to the R-matrix. LOO-CV= leave-one-hemisphere out cross-validation; LOPO-CV= leave-one-patient out cross-validation.

**Supplementary Figure S5.**
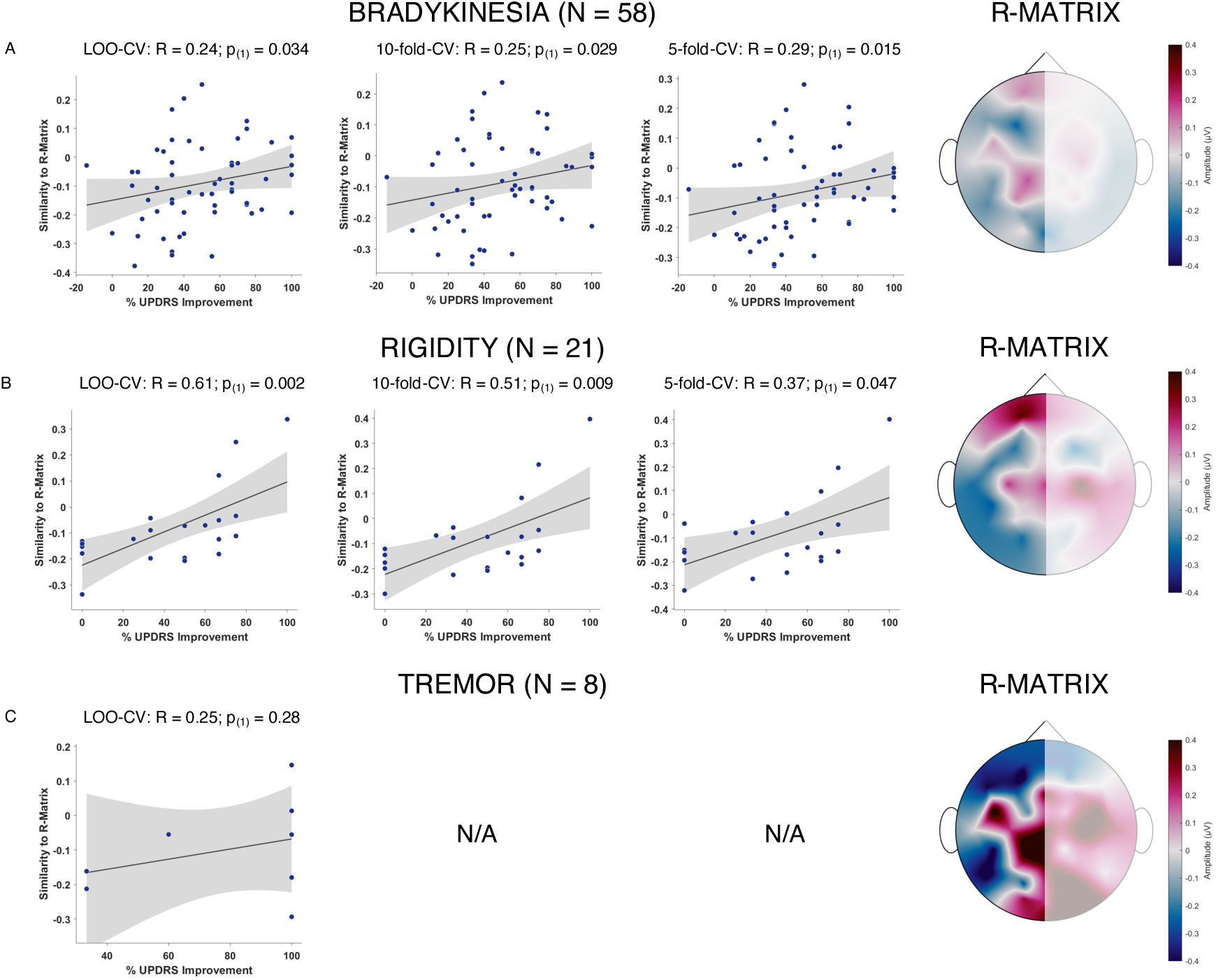
Cross-validation results using left channels for the symptom-specific R-matrices (right column shows average R-matrix topography from 15 to 40 ms). Correlation plots for each of the cross-validations using similarities to the R-matrices. Since in cross-validation experiments, only positive correlations are valid, we applied one-sided tests. For tremor, only a leave-one-out cross-validation design was tested, given the small sample size (N = 8) of patients that had at least 3 tremor points at baseline.

**Supplementary Figure S6.**
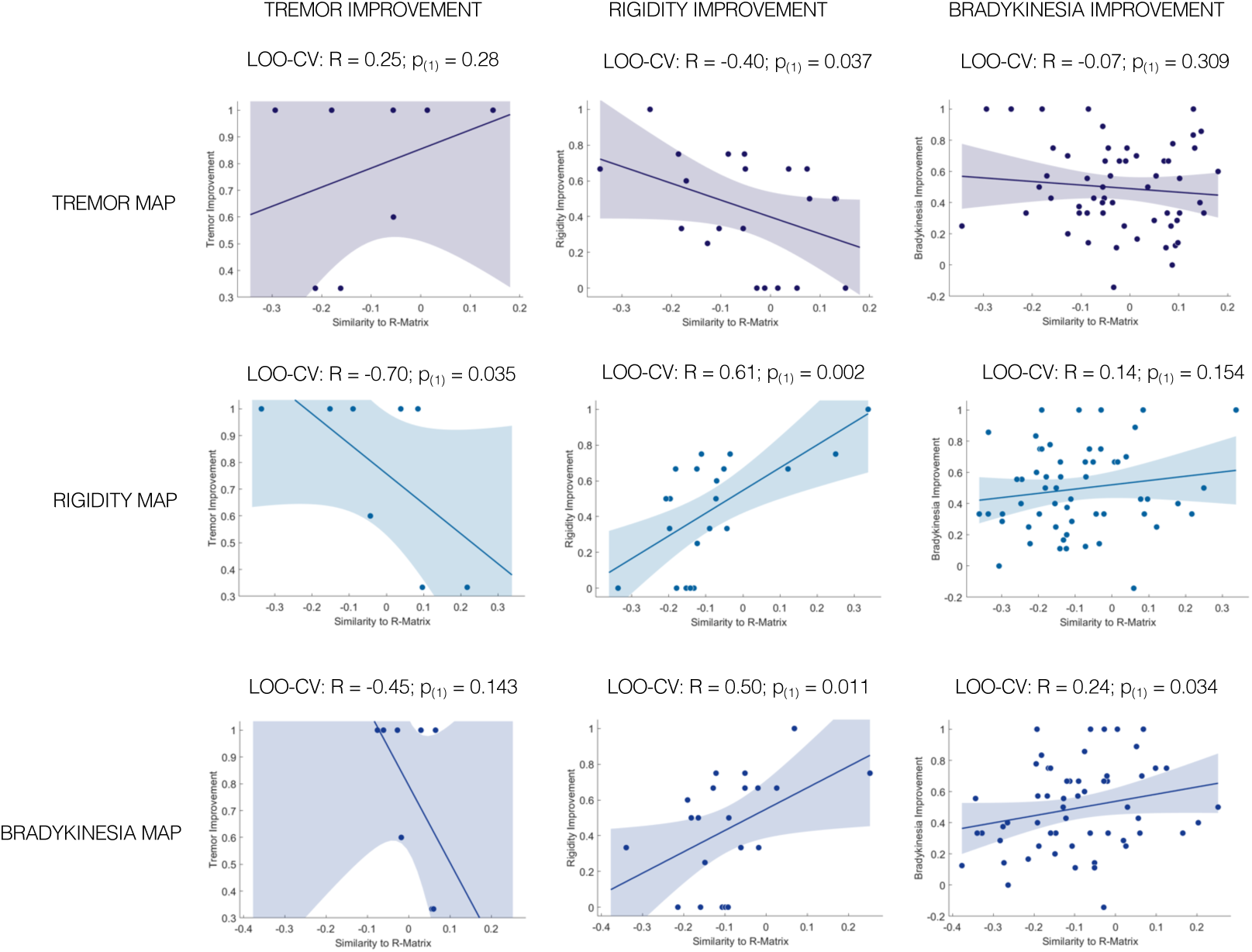
Cross-validation results using left channels for the symptom-specific R-matrices estimating each of the other as well as the same symptom improvements (each column represents the UPDRS improvements for the symptom subitems and each row the symptom R-matrices that were used to estimate the improvements). Correlation tests were one-sided. The diagonal correlation plots are identical to the leave-one-out analysis shown in supplementary figure S5.

**Supplementary Figure S7.**
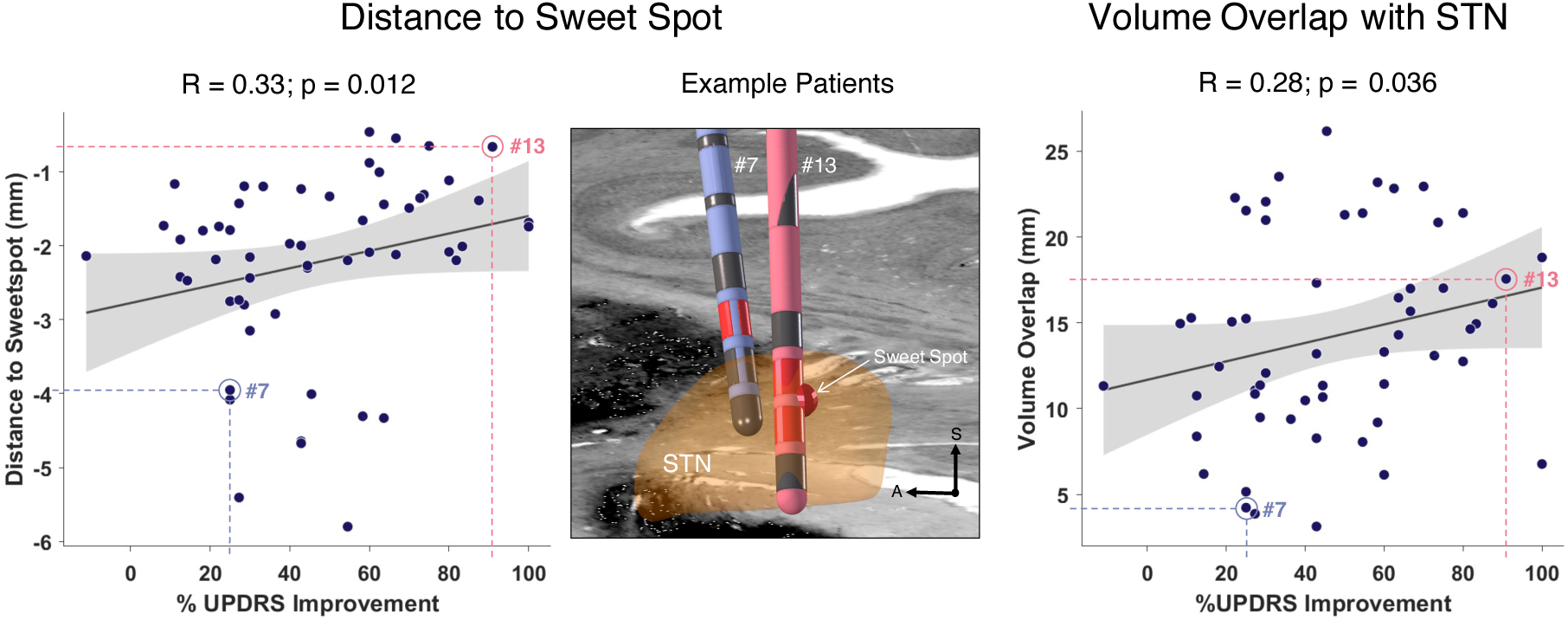
Imaging-based analysis. Correlations between distance to the motor improvement sweet spot (Caire et al.) and the e-field volume overlap with the STN (Ewert et al. 2018) with empirical UPDRS improvements. The middle panel shows two representative DBS electrodes from patient #7 and #13 in the right STN, that showed optimal (#13, red) and suboptimal (#7, blue) improvement, also reflected by the distance of the active electrode contacts (red) to the motor improvement sweet spot.

**Supplementary Figure S8.**
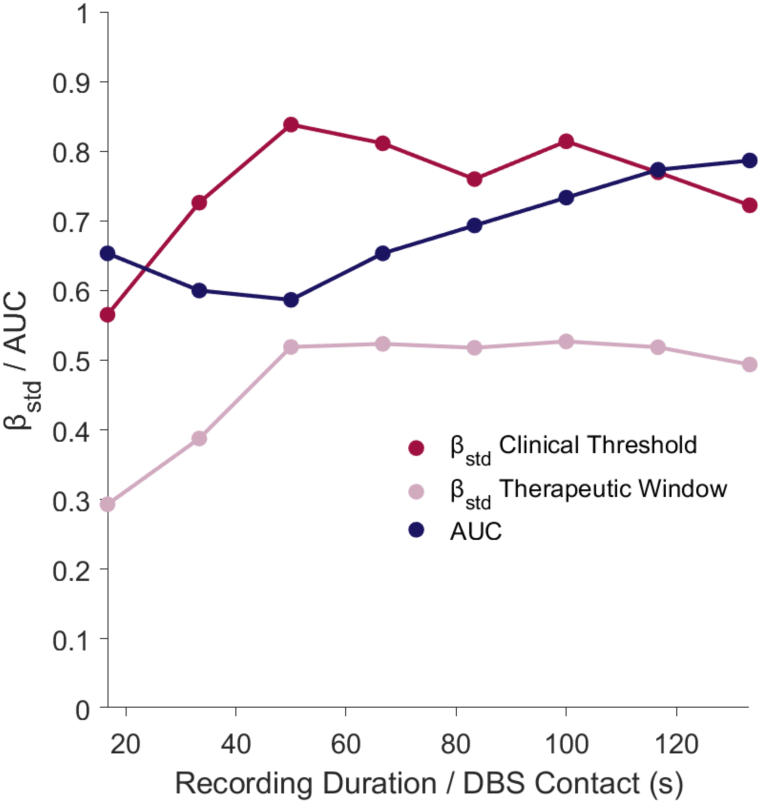
Standardized Beta coefficients (βstd) for the linear mixed effect models and area under the curve (AUC) values for receiver operating characteristics curve (ROC) analysis on the prospective validation cohort. The linear mixed effect model fits the relationship between the spatial similarity of the respective DBS contact’s EP-map to the R-matrix and the clinical effect threshold (lowest stimulation amplitude eliciting a clinical effect) as well as the therapeutic window (relative difference between the stimulation amplitude needed for a clinical effect and the amplitude eliciting side effects). The ROC analysis was conducted based on a contact selection variable (considering both, lowest clinical threshold and largest therapeutic window) and used the similarity of the respective contact’s EP-map for contact selection. Each of the three measures, beta coefficient for clinical threshold and therapeutic window as well as AUC were computed for an increasing number of stimulation epochs per recording and are shown together in one plot. At the chosen stimulation frequency of 6 Hz, the recording duration of 20 seconds is equivalent to 120 stimulation epochs and 130 s equivalent to 800 stimulation epochs. While beta coefficients were stable until a recording time of around 50 seconds (= 300 epochs), the AUC indicates a decreasing discriminative ability of the method. Only at more than 100 s (= 600 epochs) recording time, the ROC analysis indicated good discriminative abilities (AUC > 0.7) of the method.

## References

1. Deuschl, G. et al. A randomized trial of deep-brain stimulation for Parkinson’s disease. N Engl J Med 355, 896–908 (2006).

2. Roediger, J. et al. StimFit—A Data-Driven Algorithm for Automated Deep Brain Stimulation Programming. Movement Disorders 37, 574–584 (2022).

3. Picillo, M., Lozano, A. M., Kou, N., Puppi Munhoz, R. & Fasano, A. Programming Deep Brain Stimulation for Parkinson’s Disease: The Toronto Western Hospital Algorithms. Brain Stimul 9, 425–437 (2016).

4. Volkmann, J., Moro, E. & Pahwa, R. Basic algorithms for the programming of deep brain stimulation in Parkinson’s disease. Movement Disorders 21, S284–S289 (2006).

5. Boutet, A. et al. Predicting optimal deep brain stimulation parameters for Parkinson’s disease using functional MRI and machine learning. Nat Commun 12, 1–13 (2021).

6. Bahners, B. H., Spooner, R. K., Hartmann, C. J., Schnitzler, A. & Florin, E. Subthalamic stimulation evoked cortical responses relate to motor performance in Parkinson’s disease. Brain Stimul 16, 561–563 (2023).

7. Treu, S. et al. Deep brain stimulation: Imaging on a group level. Neuroimage 219, 117018 (2020).

8. Roediger, J. et al. StimFit—A data-driven algorithm for automated deep brain stimulation programming. Movement Disorders (2021) doi:10.1002/MDS.28878.

9. Rajamani, N. et al. Deep brain stimulation of symptom-specific networks in Parkinson’s disease. Nat Commun 15, 1–16 (2024).

10. Roediger, J. et al. Automated deep brain stimulation programming based on electrode location: a randomised, crossover trial using a data-driven algorithm. Lancet Digit Health 5, 59–70 (2023).

11. Ashby, P. et al. Potentials recorded at the scalp by stimulation near the human subthalamic nucleus. Clinical Neurophysiology 112, 431–437 (2001).

12. Limousin, P., Brown, P., Marsden, J., Defebvre, L. & Rothwell, J. Evoked potentials from subthalamic nucleus, internal pallidum and thalamic stimulation in Parkinsonian and postural tremor patients. Journal of Physiology **509P**, 176P–177P (1998).

13. Walker, H. C. et al. Short latency activation of cortex during clinically effective subthalamic deep brain stimulation for Parkinson’s disease. Movement Disorders 27, 864–873 (2012).

14. Baker, K. B., Montgomery Jr., E. B., Rezai, A. R., Burgess, R. & Lüders, H. O. Subthalamic nucleus deep brain stimulus evoked potentials: physiological and therapeutic implications. Movement Disorders 17, 969–983 (2002).

15. Peeters, J. et al. Towards biomarker-based optimization of deep brain stimulation in Parkinson’s disease patients. Front Neurosci 16, 1091781 (2023).

16. Spooner, R. K., Bahners, B. H., Schnitzler, A. & Florin, E. DBS-evoked cortical responses index optimal contact orientations and motor outcomes in Parkinson’s disease. NPJ Parkinsons Dis 9, 1–11 (2023).

17. Cassar, I. R. & Grill, W. M. The cortical evoked potential corresponds with deep brain stimulation efficacy in rats. J Neurophysiol 127, 1253–1268 (2022).

18. Romeo, A. et al. Cortical activation elicited by subthalamic deep brain stimulation predicts postoperative motor side effects. Neuromodulation 22, 456–464 (2019).

19. Irwin, Z. T. et al. Latency of subthalamic nucleus deep brain stimulation-evoked cortical activity as a potential biomarker for postoperative motor side effects. Clinical Neurophysiology 131, 1221–1229 (2020).

20. Bahners, B. H. et al. Deep brain stimulation device-specific artefacts in MEG recordings. Brain Stimul 17, 109–111 (2024).

21. Bahners, B. H. et al. Deep brain stimulation does not modulate auditory-motor integration of speech in Parkinson’s disease. Front Neurol 11, 655 (2020).

22. Hoffman, D., Haislip, I. & Cool, C. Estimated Time to Properly Apply Electroencephalogram (EEG) Electrodes: A Survey (P11-1.009). Neurology 102, (2024).

23. Horn, A. et al. Connectivity predicts deep brain stimulation outcome in Parkinson disease. Ann Neurol 82, 67–78 (2017).

24. Amunts, K. et al. BigBrain: An ultrahigh-resolution 3D human brain model. Science (1979) 340, 1472–1475 (2013).

25. Ewert, S. et al. Toward defining deep brain stimulation targets in MNI space: A subcortical atlas based on multimodal MRI, histology and structural connectivity. Neuroimage 170, 271–282 (2018).

26. Eusebio, A. et al. Resonance in subthalamo-cortical circuits in Parkinson’s disease. Brain 132, 2139–2150 (2009).

27. Hartmann, C. J. et al. Distinct cortical responses evoked by electrical stimulation of the thalamic ventral intermediate nucleus and of the subthalamic nucleus. Neuroimage Clin 20, 1246–1254 (2018).

28. Akram, H. et al. Subthalamic deep brain stimulation sweet spots and hyperdirect cortical connectivity in Parkinson’s disease. Neuroimage 158, 332–345 (2017).

29. Hassler, R., Riechert, T., Mundinger, F., Umbach, W. & Ganglberger, J. A. Physiological observations in stereotaxic operations in extrapyramidal motor disturbances. Brain 83, 337–350 (1960).

30. Neudorfer, C. et al. Lead-DBS v3.0: Mapping deep brain stimulation effects to local anatomy and global networks. Neuroimage 268, 119862 (2023).

31. Caire, F., Ranoux, D., Guehl, D., Burbaud, P. & Cuny, E. A systematic review of studies on anatomical position of electrode contacts used for chronic subthalamic stimulation in Parkinson’s disease. Acta Neurochir (Wien) 155, 1647–1654 (2013).

32. Horn, A. et al. Lead-DBS v2: Towards a comprehensive pipeline for deep brain stimulation imaging. Neuroimage 184, 293–316 (2019).

33. Bahners, B. H. et al. Electrophysiological characterization of the hyperdirect pathway and its functional relevance for subthalamic deep brain stimulation. Exp Neurol 352, 114031 (2022).

34. Miocinovic, S. et al. Cortical potentials evoked by subthalamic stimulation demonstrate a short latency hyperdirect pathway in humans. The Journal of Neuroscience 38, 9129–9141 (2018).

35. Jorge, A. et al. Hyperdirect connectivity of opercular speech network to the subthalamic nucleus. Cell Rep 38, 110477 (2022).

36. Steiner, L. A. et al. Neural signatures of indirect pathway activity during subthalamic stimulation in Parkinson’s disease. Nat Commun 15, 1–13 (2024).

37. Spooner, R. K., Hizli, B. J., Bahners, B. H., Schnitzler, A. & Florin, E. Modulation of DBS-induced cortical responses and movement by the directionality and magnitude of current administered. npj Parkinson’s Disease 2024 10:1 **10**, 1–10 (2024).

38. Hollunder, B. et al. Toward personalized medicine in connectomic deep brain stimulation. Prog Neurobiol 210, 102211 (2022).

39. Bhanpuri, N. H. et al. Deep brain stimulation evoked potentials may relate to clinical benefit in childhood dystonia. Brain Stimul 7, 718–726 (2014).

40. Waters, A. C. et al. Test–retest reliability of a stimulation-locked evoked response to deep brain stimulation in subcallosal cingulate for treatment resistant depression. Hum Brain Mapp 39, 4844–4856 (2018).

41. Seas, A. et al. Subcallosal cingulate deep brain stimulation evokes two distinct cortical responses via differential white matter activation. Proc Natl Acad Sci U S A 121, e2314918121 (2024).

42. Dähne, S. et al. SPoC: A novel framework for relating the amplitude of neuronal oscillations to behaviorally relevant parameters. Neuroimage 86, 111–122 (2014).

43. Jost, S. T. et al. Levodopa Dose Equivalency in Parkinson’s Disease: Updated Systematic Review and Proposals. Movement Disorders 38, 1236–1252 (2023).

44. Yu, A. B. & Hairston, W. D. Open EEG Phantom. (2019) doi:10.17605/OSF.IO/QRKA2.

45. Tadel, F., Baillet, S., Mosher, J. C., Pantazis, D. & Leahy, R. M. Brainstorm: A user-friendly application for MEG/EEG analysis. Comput Intell Neurosci 2011, (2011).

46. Hollunder, B. et al. Mapping dysfunctional circuits in the frontal cortex using deep brain stimulation. Nat Neurosci 27, 573–586 (2024).

47. Oxenford, S. et al. WarpDrive: Improving spatial normalization using manual refinements. Med Image Anal 91, 103041 (2024).

48. Husch, A., V. Petersen, M., Gemmar, P., Goncalves, J. & Hertel, F. PaCER - A fully automated method for electrode trajectory and contact reconstruction in deep brain stimulation. Neuroimage Clin 17, 80–89 (2018).

49. Horn, A. & Kühn, A. A. Lead-DBS: A toolbox for deep brain stimulation electrode localizations and visualizations. Neuroimage 107, 127–135 (2015).

50. Butenko, K., Bahls, C., Schröder, M., Köhling, R. & Van Rienen, U. OSS-DBS: Open-source simulation platform for deep brain stimulation with a comprehensive automated modeling. PLoS Comput Biol 16, e1008023 (2020).

